# From sequence data to patient result: a solution for HIV drug resistance genotyping with Exatype, end to end software for Pol-HIV-1 Sequence analysis and patient HIV drug resistance result generation

**DOI:** 10.1101/2020.01.29.20019448

**Authors:** Leonard King’wara, Muthoni Karanja, Catherine Ngugi, Geoffrey Kangogo, Kipkerich Bera, Maureen Kimani, Nancy Bowen, Dorcas Abuya, Violet Oramisi, Irene Mukui

## Abstract

**Introduction:** With the rapid scale-up of antiretroviral therapy (ART) to treat HIV infection, there are ongoing concerns regarding probable emergence and transmission of HIV drug resistance (HIVDR) mutations. This scale-up has to lead to an increased need for routine HIVDR testing to inform the clinical decision on a regimen switch. Although the majority of wet laboratory processes are standardized, slow, labor-intensive data transfer and subjective manual sequence interpretation steps are still required to finalize and release patient results. We thus set out to validate the applicability of a software package to generate HIVDR patient results from raw sequence data independently.

**Methods:** We assessed the performance characteristics of Hyrax Bioscience’s Exatype (a sequence data to patient result, fully automated sequence analysis software, which consolidates RECall, MEGA X and the Stanford HIV database) against the standard method (RECall and Stanford database). Exatype is a web-based HIV Drug resistance bioinformatic pipeline available at sanger.exatype.com. To validate the exatype, we used a test set of 135 remnant HIV viral load samples at the National HIV Reference Laboratory (NHRL).

**Result:** We analyzed, and successfully generated results of 126 sequences out of 135 specimens by both standard and Exatype software. Result production using Exatype required minimal hands-on time in comparison to the standard (6 computation-hours using the standard method versus 1.5 Exatype computation-hours). Concordance between the two systems was 99.8% for 311,227 bases compared. 99.7 % of the 0.2% discordant bases, were attributed to nucleotide mixtures as a result of the sequence editing in Recall. Both methods identified similar (99.1%) critical antiretroviral resistance-associated mutations resulting in a 99.2% concordance of resistance susceptibility interpretations. Base calling comparison between the two methods had Cohen’s kappa (0.97 to 0.99) implying an almost perfect agreement with minimal base calling variation. On a predefined dataset, RECall editing displayed the highest probability to score mixtures accurately one vs. 0.71 and the lowest chance to inaccurately assign mixtures to pure nucleotides (0.002–0.0008). This advantage is attributable to the manual sequence editing in RECall.

**Conclusion:** The reduction in hands-on time needed is a benefit when using the Exatype HIV DR sequence analysis platform and result generation tool. There is a minimal difference in base calling between Exatype and standard methods. Although the discrepancy has minimal impact on drug resistance interpretation, allowance of sequence editing in Exatype as RECall can significantly improve its performance.

## Introduction

Human immunodeficiency virus (HIV) drug resistance testing (DRT) has been used by WHO to guide policies relating to antiretroviral treatment (ART) dispensation at an individualized level in clinical practice as well as the public health recommendations for antiretroviral therapy regimens in various populations^1,2^. Drug resistance testing identifies mutations within the viral genome that confer resistance to the patient regimen, thus allowing healthcare workers to optimize patient treatment, increasing the chance of successful virologic suppression. Furthermore, implementing platforms for drug resistance surveillance at the population level^3,4^ can help minimize the use of ineffective drugs, improving population-wide treatment outcomes, and reducing the risk of transmitted HIV drug resistance^5–7^. The ThermoFisher in-house genotyping procedure is one of the methods widely used for HIV drug resistance testing.

The HIV DRT wet laboratory processes includes several steps such as viral RNA extraction using plasma or dry blood spot (DBS) sample type, reverse transcriptase-polymerase chain reaction PCR (RT-PCR) amplification, nested PCR, gel documentation, nested PCR product clean-up, cycle sequencing, cycle sequencing product clean up and finally population-based (bulk) sequencing^8,9^. Several sequencing primers depending on the laboratory method, are required during the sequencing step to ensure complete bidirectional coverage over the entire length of the HIV-1 pol region of interest. In our laboratory, a laboratory specialist then assesses the quality of the sequences using an Applied Biosystems (ABI) sequence scanner before transferring ABI sequence trace files from the genetic analyzer to a disc or flash drive. DNA sequence reads from each specimen are then separately assembled into a contiguous consensus sequence in FASTA format by use of RECall analysis software (web or standalone). Sequence scanner and MEGA X is used to assess the quality of the FASTA file for contamination check using phylogenetic analysis method, and eventually transfer to the HIVDB Stanford database for mutation interpretation. These steps require considerable hands-on time as well as a highly trained technician. These steps can be challenging and time-consuming in a busy HIV DRT laboratory that is processing more than 300 samples per week with limited human resources.

Despite the number of HIV DR laboratories in resource-limited settings moving to RECall as standard software for contig assembly, resulting in the standardization of result reporting, resistance mutation reporting still varies in some cases between laboratories, even between identical samples ^10,11^. Most of these inter-laboratory discrepancies come from differences in sample preparation procedures (e.g., extraction procedures, primer choice, quality assurance adherence, clean-up processes, or stochastic variation). However, some are still as a result of the change introduced by technicians as they subjectively review the assembled sequences ^12,13^. With the introduction of test and treat policies resulting in rapid ART initiation among those newly diagnosed with HIV^14^, most drug-resistant HIV variants are present at low frequencies in clinical isolates. Thus accurate identification of nucleotide “mixtures” (positions having two or more nucleotides) is required, especially for DR surveillance^15–17^. Limited laboratory specialist capabilities and experiences in low-level nucleotide mixtures identification could thus result in clinically relevant drug resistance mutations being missed^10,18^. Even though, standardization of laboratory Quality practices and protocols among the WHO-accredited laboratories has been instituted by external quality assurance program ^18^, the process does not capture HIV DR testing laboratories out of the WHO HIVResNet even though these laboratories do support the patient diagnosis. Also, despite these QA programs being in place, the impact of erroneous results due to subjective sequence interpretation on patient care is difficult to ascertain.

Implementation of automated sequence analysis and result reporting tool would enable objective and consistent interpretation and reporting of HIV genotype data and provide considerable practical advantages to clinicians, patients, and other healthcare workers utilizing the results for patient management. Most notably, it would improve result processing speed and significantly decrease human resource needs that would be apparent in the implementation of routine DR testing in resource-limited settings. The National HIV Reference laboratory in Kenya has thus validated a bioinformatics software tool, Exatype, that has the capabilities to address these challenges. Exatype consolidates the WHO-adopted processes for HIV DR genotyping into a single step - contig assembly, mutation calling, and drug-resistance interpretation are all automated. Specifically, Exatype includes the RECall software to interpret and analyze chromatograms and the Stanford HIVDB drug resistance algorithm for drug-resistance interpretation. Besides, it contains genetic distance analysis that allows for the detection of contamination. As an automated process, Exatype is to support HIV DR testing laboratories with a heavy workload. It combines the functionalities of RECall, Stanford HIV drug resistance database (HIVDB) and, MEGA X programs and is available at sanger.exatype.com.

In this paper, we present field validation results for automated Exatype analysis and reporting of HIV DR results.

## Materials and methods

### Laboratory methods

HIV drug resistance testing was performed at the National HIV Reference laboratory in Kenya. Using 1000 copies/ml program guidelines cut off for viral suppression as the test kit sensitivity limit, we picked remnant samples from the HIV RNA measurement section after HIV-1 viral load testing^20^. We performed HIV genotypic resistance testing on 135 remnant patient samples. We did plasma virus extraction using the ThermoFisher Kingfisher flex platform followed by one-step RT-PCR, denaturing of amplicons, and finally, a nested second-round PCR. For QA purposes, we assessed the PCR product on a gel. The cleanup procedure used Exosap before proceeding to cycle sequencing. Also, sequence product purification used x-terminator. An ABI 3730xl performed direct bi-directional sequencing encompassing HIV-1 protease (PR) and the first 296 codons of reverse transcriptase (RT). Sequencing Analysis v 5.2 (ABI) assisted in reading the chromatograms. For quality assurance, nucleotide mixtures (positions containing two or more nucleotides, with the minor peak height being ≥20% of the significant peak height, were marked with ABI 3730xl data collection software v 3.30.

### Standard analysis procedure

After the necessary sequence QA procedures using a sequence scanner, a laboratory specialist assembles the sequence trace files for each sample to generate a consensus sequence using standalone RECall software. This software assists the specialist by highlighting areas of conflicts as nucleotide positions with mixture and where overlapping sequence positions do not have the same base call (20% threshold). N is used to mark undistinguished regions of the sequence chain. The laboratory specialist then visually inspects each sequence, stopping at each conflict and making manual edits where necessary. This verification is to ensure that any variations are verified. The generated consensus sequence for each sample is then subjected to MEGA X for contamination check analysis and later to Stanford HIVDB to create patients’ results. In addition to the 135 patient specimens, we included 40 EQA dry panels from the WHO ResNet Lab group to ensure that the study conforms to the Clinical and Laboratory Standard Institute CLSI guidelines on laboratory method validations.

### Besides the standard method, we used Exatype

to reanalyze and generate results from 3730xl ABI trace files without sequence editing. Similarly, to RECall, overlapping peaks represents “mixed or ambiguous” bases. The location of the primary peak (called base) and the most significant secondary peak (uncalled base) in the trace file are determined by phred. It then aligns the peak positions to their corresponding locations in the. ab1 data as most primary and secondary peaks often offset. Poor sequence quality regions at the beginning and end of each fragment are then automatically identified and trimmed. All chromatograms (. ab1) were submitted to Exatype and processed without any human intervention, using a standard laptop (Asus-i3 660 3.33-GHz CPU, 3 GB RAM, Windows 2010).

### Exatype nucleotide mixture calling and “marking” of potentially problematic bases

The essential feature of Exatype is its consolidated workflow, where no file transfer between separate software programs is necessary. The contig assembly and FASTA file generation (by implementing RECall) and the subsequent interpretation by Stanford HIVDB is done automatically, without any editing or file transfer to MEGA X for contamination check or Stanford HIVDB for result generation. Following the assembly and alignment step, mixtures categorization is based on the quality and area under the curve of the called and uncalled base as determined by Phred using a built-in RECall software. Configuration for RECall, within Exatype, variables that guide its mixture calling for clinical drug resistance testing at the National HIV Reference Laboratory (NHRL) are listed in **Table 2**. The examination of each position in the sequence alignment sequentially and the samples that require manual editing are marked.

**Table 1:** Distribution of the HIV-1 subtype in the samples used for validation.

|  |  |
| --- | --- |
| **Subtype A** | 54(43%) |
| **Subtype D** | 31(25%) |
| **Subtype C** | 14(11%) |
| **Subtype G** | 8(6%) |
| **Subtype AD** | 7(6%) |
| **Subtype AC** | 2(2%) |
| **Subtype AG** | 2(2%) |
| **Subtype CRF01_AE** | 8(6%) |
| **VL < 1000 cp/ml** | 34(27%) |
| **VL > 1000 cp/ml** | 92(73%) |
| Therapy naive | 9(7%) |
| Therapy experience | 103(82%) |
| Therapy unknown | 14(11%) |

**Table 2:** Configuration variables for nucleotide mixture calling and base “marking” for clinical drug resistance genotyping^22,23^.

| Parameter | Value | Interpretation |
| --- | --- | --- |
| **Quality censoring cutoff** | $< 10$ | Phred quality scores cut off for excluding bases during assembly. |
| **Mixture area (%)** | $\geq 20$ | The area of the uncalled peak must be at least 20% of the called peak area. If $\geq 50\%$ of the reads pass this threshold, then a mixture is called. |
| **Mark area (%)** | $\geq 15$ | The area of the uncalled peak must have at least 17.5% of the called peak area. If $\geq 50\%$ of the reads pass this threshold, then a mark is made. |
| **Mark average quality cut-off**<br>**phred score Additional marks** | $< 20$ | If the average quality of the base across all reads is below the cutoff, then a mark is made. Insertions, deletions, and single primer coverage are also marked. |

### Exatype pass-fail criteria at the laboratory level

We performed quality checks on every sample trace file to ensure that the sequence was acceptable. Table 2 lists the sequence rejection criteria. Once the trace file is uploaded to Exatype, and it passes the RECall specified internal quality control checks, it automatically generates sample results and corresponding FASTA files. At present, the software requires double primer coverage over the entire sequence length. We included only analyses that passed the Exatype-implemented quality control criteria in this study.

### Subtyping and phylogenetic analysis

One hundred and twenty-six samples were successfully extracted, amplified for the *gag* gene in a nested PCR, and sequenced. Generated sequences (approximately covering 460–470 bp of p24 and p7 region of *gag* gene, NT 1577–2040, HXB2) were then used for sequence alignment using RECall and to construct the phylogenetic trees using the neighbor-joining method with PAUP. Alignment of generated sequences with Los Alamos database reference sequences revealed that 54 (43%),31 (25%),14 (11%), and 8 (6%) of the 126 specimens are subtype A, D, C, and G respectively. *Simplot* analysis revealed a few recombinant types in our study samples: 7 (6%) being AD, 2 (2%) AC, 2 (2%) AG and 8 (6%) CRF01_AE (Table 1).

### Data analyses

We compared the consensus sequences and results generated by the standard method and Exatype. Speed, concordance of base calls, and results were used to asses the performance of Exatype. Partial nucleotide discordance is when one methodology reported a nucleotide mixture, and the other reported one of the mixture’s components (e.g., RECall reported Y and Exatype reported C). Complete nucleotide discordance is when the two-analysis method used, indicate different nucleotide at the same position for the same sample (e.g., RECall reported T and Exatype indicate C). Similarly, this can occur in a mixture when nucleotide called by one method is different from the other (e.g., RECall reported G and Exatype indicate Y).

We also compared the analysis of specific antiretroviral drug resistance mutation positions as defined by International AIDS Society (USA table) on key resistance mutations. We processed 126 samples on Stanford HIV drug resistance genotyping Web service Sierra (algorithm version 8.8 [http://hivdb.stanford.edu/pages/algs/sierra_sequence.html]; Stanford University, Stanford, CA) to infer antiretroviral drug susceptibilities in RECall analyzed PR-RT nucleotide sequences. ANRS version 27, HIVDB version 8.9-1, and REGA version 8.0.2 reanalyzed the samples. For, 10% false-positive rate (FPR) or 20% FPR cut-off, we used G2P Geno2Pheno for prediction in the coreceptor.

### Ethics statement

Amref Health Africa Research Ethics Committee approved the study (Ref No, 4562). We used the principles of the international Declaration of Helsinki 2013 and Good clinical laboratory practices to conduct the research. The study used a waiver of consent to conduct analyses on the remnant HIV viral load samples. Patients were receiving clinically significant results.

## Results

RECall was able to generate a consensus sequence for 9.8% (132/135) of the pol experiments, whereas Exatype was successful in 93.3% (126/135) of the tests (**Table 4**). Of these, 126 (93.3%) met the default Exatype and RECall acceptability criteria after automated processing. Inadequate double primer coverage over the entire sequence length was the primary reason for failure. For the standard analysis using a standard Laptop (ASUS-i3 660 3.33-GHz CPU, 3 GB RAM, Windows XP), we performed RECall base calling, assembly, contamination check using MEGA X and alignment in less than 4 hours, with human sequence edit review. We then proceeded and used Stanford HIVDB to generate patient results in one hour.

**Table 3:** Criteria used by RECall for rejecting a sequence ^9,18^.

| Failure category | Description |
| --- | --- |
| **Stop codon** | Any unambiguous stop codon (TGA, TAA, or TAG) |
| **Bad inserts** | An insertion relative to the reference sequence that is not a multiple of three |
|  | *bases, resulting in a frameshift* |
| **Bad deletion** | *A deletion relative to the reference sequence that is not a multiple of three bases, resulting in a frameshift* |
| **Too many mixtures** | *>3.5% of nucleotides sequences called as mixtures* |
| **N count** | *>=5 Ns (any base) in the sequence* |
| **Mark count** | *>=100 positions marked as being potentially problematic* |
| **Single coverage** | *>3 consecutive bases of single-read coverage with phred scores of 40* |
| **Low quality** | *Any section where the quality of all coverage is too low to make a call* |

**Table 4:** Performance in generating consensus pol sequences for HIV-1 samples by the different editing approaches.

| Editing method | Results | No Results | Total |
| --- | --- | --- | --- |
| Exatype | 126 (93%) | 9 (7%) | 135 |
| Standard analysis Procedure | 132 (98%) | 3 (2%) | 135 |

In contrast, we did the entire analysis, QA contamination report generation, and patient result generation in Exatype on the same laptop within one hour. The longer time in the standard software pipeline is attributed from the sequence review and edits before exporting the contig into a different software MEGA X for QA analysis and Stanford HIVDB for patient result generation. All the steps are performed simultaneously in Exatype.

### Nucleic acid sequence concordance between Exatype and Standard analysis procedure

Within 311,227 analyzed bases, there was 99.8% overall agreement in base calling between Exatype and the gold standard. There was 99.6% complete sequence concordance within 311,227 nucleotide positions, as indicated in **Fig. 1**. Of the 311 discordant nucleotides, 308 (99%) were “partially discordant” (mixtures called by one method but not the other), while 3 (1%) were wholly discordant. 76.5% (238 of 311) of the partially different bases comprised of nucleotide pairs as a result from transitions (R A/G, Y C/T) rather than transversions (K G/T, M A/C, S C/G, W A/T).

**Figure 1.**
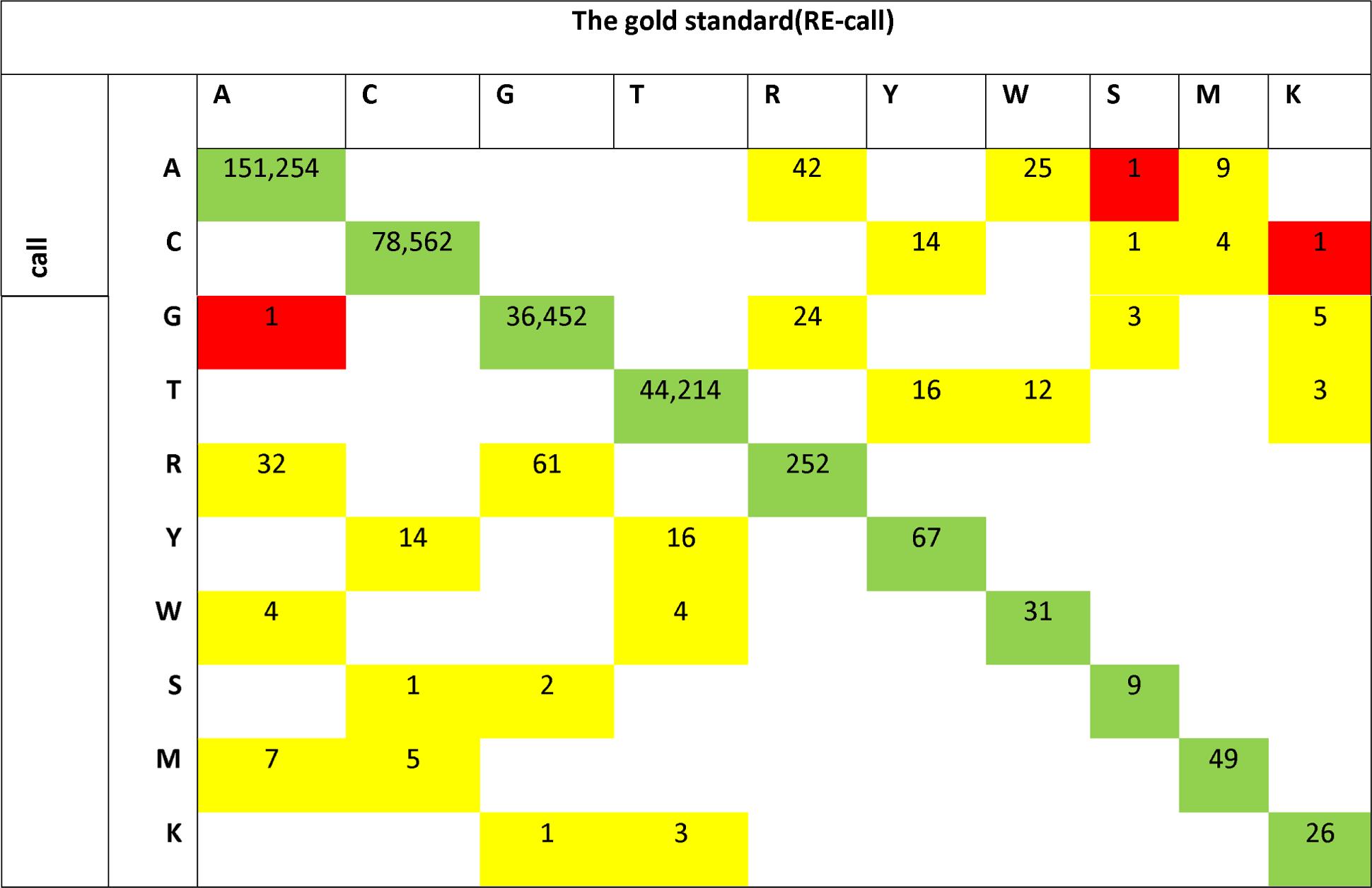
Concordant and discordant nucleotide base call in sequences analyzed by Gold standard and by Exatype. Matrices depict the frequencies of nucleotides called by Exatype (vertical axis) and by Standard (horizontal axis). Green highlight implies a concordant base call. Partially discordant base calls (implies mixtures called by one method but not the other) is in yellow highlight. Entirely discordant base calls are in red highlight. Zero is represented by blank cells. International Union of Biochemistry and Molecular Biology ambiguity codes are as follows; R A/G, Y C/T, W A/T, M A/C, K G/T, S G/C, B C/G/T, D A/G/T, H A/C/T, and V A/C/G. Overall, 99.8% concordance was observed for 311,227 bases compared.

Distribution of discordant positions between the transitions, transversions, and a combination of both was relatively the same (*n* 11, 6, and 5, respectively), as indicated in **Fig. 1**. 1.2% of nucleotide mixtures detected on all bases. Overall, the standard method called a marginally more significant number of mixtures (1193 standard method -called mixtures [1.08%] and 1181 Exatype-called mixtures [1.05%]; *P* 0.6).

### Amino acid sequence concordance between Gold standard and Exatype interpretations

the 311 discordant nucleotide positions resulted in 284 discordant codons. 114 (40.1%) of these, produced Nonsynonymous substitutions between the standard and Exatype method at the sequence to amino acid translation level. 278 (97.8%) were partial amino acid discordances (sharing at least one amino acid between the two interpretations), while only 6 (2.2%) were complete amino acid differences.

In general, the gold standard and Exatype sequence review identified 97 “key” antiretroviral drug resistance mutations^24^, as either complete amino acid substitutions or as part of mixtures. The two methods agreed for 123 cases. The Exatype identified one resistance mutation (E35D) that the gold standard did not, while the gold standard identified 2(K55R and R57K) that Exatype did not. This variation in resistant mutation identification affected two patient results though none of the three mutations has clinical significance

From the HIV-1 RNA measurement remnant samples, 93.3% (126/135) of the pol HIV-1 had a consensus NT sequences available and generated by both Exatype and RECall (Table 5). In total, 86.5% (109/126) of the PR, 74.6% (94/126) of the RT sequences were fully concordant at the NT level. At the AA level, 81% (102/126) of the PR, 51.1% (72/126) of the RT samples were the same. The differences in concordance between the different regions were attributed to the difference in coverage length and were less pronounced when normalized.

**Table 5:**
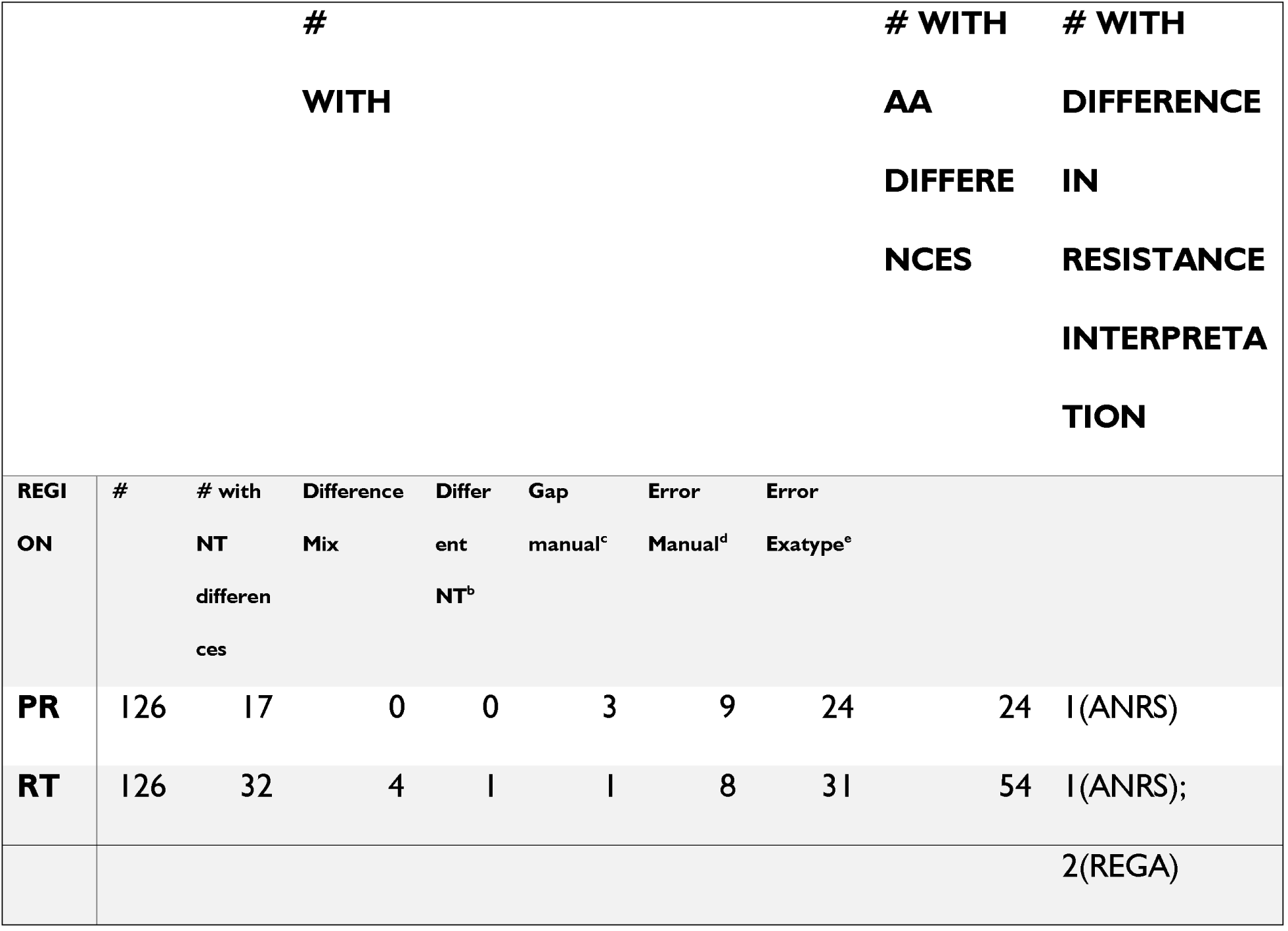
Differences in Gold standard and Exatype editing of HIV-1 pol sequences from clinical samples and impact on drug resistance interpretation. We considered sequences that passed both Exatype and Gold standard editing. #, number of samples; NT, nucleotide; AA, amino acid; genotypic drug resistance interpretation systems: ANRS version 27, HIVDB version 8.9-1, and REGA version 8.0.2. We used G2P Geno2Pheno for prediction in coreceptor use when cut-off of 10% false-positive rate (FPR) or 20% FPR. a) Number of samples with mixtures scored differently by the two approaches b) Number of samples with pure nucleotides scored separately by the two approaches c) Number of samples with parts of sequences that were not analyzable as judged by the editor d) The number of samples containing differences between Recall and Exatype editing due to manual editing. e) Number of samples containing differences between Exatype and Recall editing due to errors made during automatic editing in Exatype

For each discordant NT call, the chromatograms were manually reviewed by a second laboratory specialist to verify whether the differences resulted from an erroneous call in the automatic or manual editing process. For both editing approaches, incorrect calls were observed, i.e., in 24 vs. nine samples for PR, 31 vs. 1 for RT (Table 5). Only 1 RT nucleotide was different between the manually and automatically edited sequences. In both instances, differences result from mistakes made during manual editing. The operator trimmed the five ends of PR in 3 samples and one sample for RT, but these parts were still completely analyzed by RECall and not Exatype. Additionally, some of the erroneous calls in Exatype were because this tool does not allow sequence editing.

85% (22 + 12 = 34)/40) of the EQA dry panels (These are FASTA files shared by the WHO to all the WHO accredited lab for competency assessment of staff in sequence editing) from WHO had a consensus sequence using Recall, while for the Exatype, it was 80% (32/40) (**Table 6**). For each dry panel, a reference sequence sent by WHO was considered as the accurate results, and was calculated based on the consensus results of all participants within the WHO ResNet Lab (∼52 participants). We further reviewed each discordant NT call to find out whether the difference resulted from a missed mixture, a false mixture, or a different NT or mixture (Table 6). Both Recall and Exatype are comparable in terms of detecting mixtures with both almost having a similar score on the mixtures that were not present in the reference sequence (**Table 6**).

**Table 6:**
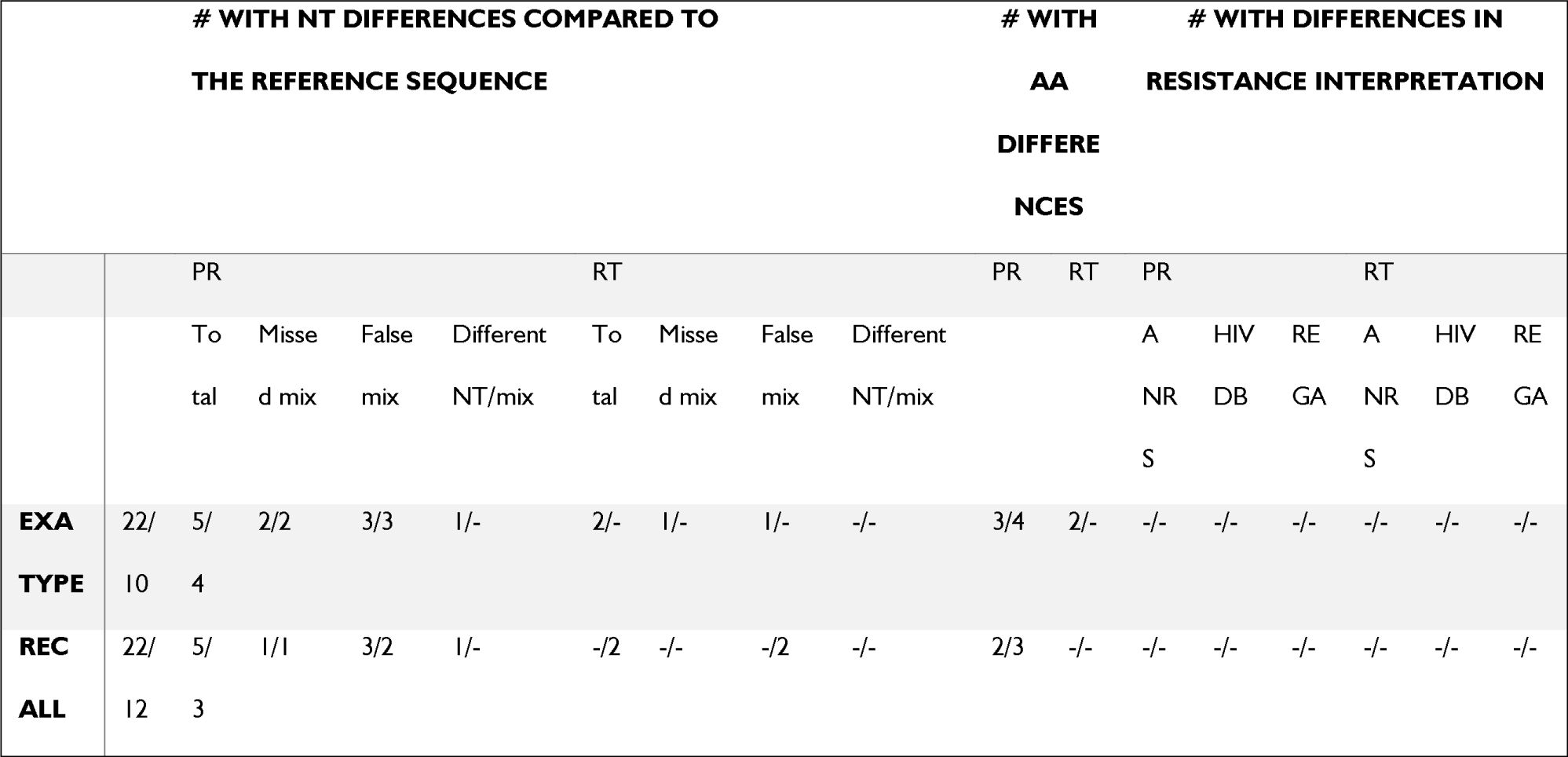
Differences in RECall and Exatype editing of HIV-1 pol sequences from all EQA samples and impact on drug resistance interpretation. This analyses were confined to drug resistance positions (PR: 10, 20, 24, 30, 32, 33, 36, 46, 47, 48, 50, 53, 54, 63, 71, 73, 77, 82, 84, 88, 90; RT: 41, 62, 65, 67, 69ins, 69, 70, 74, 75, 77, 100, 103, 106, 108, 115, 116, 151, 181, 184, 188, 190, 210, 215, 219, 225). #, number of samples; PR, protease; RT, reverse transcriptase; NT, nucleotide; AA, amino acid; genotypic drug resistance interpretation systems: ANRS version 27, HIVDB version 8.9-1 and REGA version 8.0.2. The number of sequences that passed Exatype and RECall editing are before the slash. Number of sequences that did not pass either of the two approaches are behind the slash I. The number of samples with mixtures present in the reference sequence, but not scored by the editing approach (pure wild-type or mutant NT). II. The number of samples with mixtures scored by the editing approach that was not present according to the reference sequence (pure wild-type or mutant NT). III. Number of samples with mixtures and pure nucleotides scored differently by the editing approach and the reference sequence

At the NT level, the percentage of sequences without differences compared to the reference sequence is the slightly lower for Exatype editing, which is 75% and 94% for PR and RT, respectively vs. 82% and 94% for RECall editing (Table 7). Using Recall, 0.43% of the PR and 0.04 % of the RT nucleotides were discordant with the reference sequence, in contrast to 0.88% of the PR and 0.17% of the RT nucleotides using Exatype which was markedly higher. The same tendency observed at the AA level (Table 7). We then assessed for editing approach, the probability P(M_e_|M_r_) that a mixture scored if the mixture was present in the reference sequence and the probability P(M_e_|P_r_) that a mixture scored yet it was a pure nucleotide sequence.

**Table 7:** Comparison of RECall, Exatype editing of WHO dry sample EQA panel with the reference sequence at NT and AA level. To meet the CLSI guidelines of 40% reference panels being EQA standards, we included dry panels from the WHO ResNet group. #, number of; AA, amino acids; NT, nucleotides; M_e_, mixtures present in the results of the editing approach; M_r_, mixtures present in the reference sequences; M_e_ ∩ M_r_, mixtures present in the reference sequences that scored as a mixture by the editing approach; P_r_, pure nucleotides present in the reference sequences; P(M_e_|M_r_), the probability that a mixture scored if present in the reference sequence; M_e_ ∩P_r_, pure nucleotides in the reference sequences that scored as a mixture by the editing approach; P(M_e_|P_r_), the probability that a mixture scored if no mixture was present in the reference sequence

|  | RECALL |  | EXATYPE |  |
| --- | --- | --- | --- | --- |
|  | PR | RT | PR | RT |
| # SEQUENCES WITHOUT NT DIFFERENCES | 28/34(82%) | 32/34(94%) | 24/32(75%) | 30/32(94%) |
| # SEQUENCES WITHOUT AA DIFFERENCES | 30/34(88%) | 34/34(100%) | 26/32(81%) | 30/32(94%) |
| # NT DIFFERENCES/TOTAL # NT | 9/2112(0.43%) | 1/2562(0.04%) | 18/2023(0.88%) | 4/2400(0.17%) |
| # AA DIFFERENCES/TOTAL # AA | 5/724(0.72%) | 0/912(0%) | 10/675(1.48%) | 2/800(0.25) |
| # $M_E \cap M_R$ | 18 | 18 | 12 | 8 |
| # $M_R$ | 21 | 19 | 16 | 11 |
| $P(M_E \vert M_R)$ | 0.83 | 1 | 0.7 | 0.85 |
| # $M_E \cap P_R$ | 7 | 2 | 8 | 1 |
| # $P_R$ | 2081 | 2679 | 1999 | 2381 |
| $P(M_E \vert P_R)$ | 0.002 | 0.0008 | 0.004 | 0.0004 |

In the remnant HIV-1 RNA samples, the majority of samples for which at least one of the editing approaches was able to generate a consensus NT sequence were interpreted as susceptible to most PI, NRTI, and NNRTI and scored as using the CCR5 coreceptor (Table 8) solely. Also, much more extensive drug resistance profiles observed in the WHO dry panel as compared to the clinical dataset (Table 8).

**Table 8:** Number of samples displaying (intermediate) resistance to different drug classes, according to ANRS, HIVDB, REGA, and Geno2Pheno. For the HIV-1 RNA remnant dataset, we included only the sequences that passed for both RECall, and Exatype editing. In contrast, we included resistance information of all reference sequences for the WHO dry panel dataset. FPR, false-positive rate; RTI, reverse transcriptase inhibitor; PI, protease inhibitor; genotypic drug resistance interpretation systems: ANRS version 27, HIVDB version 8.9-1 for the clinical dataset and HIVDB version 8.9-1 for the EQA dataset, and REGA version 8.0.2, G2P Geno2Pheno

| DATA SET | ACCORDIN<br>G TO | ANRS |  | HIVDB |  | REGA |  | G2P | G2P FPR |
| --- | --- | --- | --- | --- | --- | --- | --- | --- | --- |
|  |  |  |  |  |  |  |  | FPR 10% | 20% |
| CLINICAL |  | PI | RTI | PI | RTI | PI | RTI | CCR5 | CCR5 |
|  |  |  |  |  |  |  |  | antagonists | antagonists |
|  | Exatype | 82/126 | 51/126 | 33/126 | 42/126 | 34/126 | 41/126 | 49/126 | 50/126 |
|  | FASTA file | (65%) | (41%) | (26%) | (33%) | (27%) | (33%) | (39%) | (40%) |
|  | RECall | 81/126 | 52/126 | 33/126 | 42/126 | 34/126 | 43/126 | 48/126 | 50/126 |
|  | FASTA file | (64%) | (41%) | (26%) | (33%) | (27%) | (34%) | (38%) | (40%) |
| WHO DRY | Reference | 18/40 | 20/40 | 18/40 | 20/40 | 17/40 | 20/40 |  |  |
| PANEL |  | (45%) | (50%) | (45%) | (50%) | (43%) | (50%) |  |  |

## Discussion

The study evaluated the performance characteristics of the Exatype sequence analysis and result generation tool developed by Hyrax Biosciences, the ability to accurately analyze and interpret ABI sequence data into patient results. Exatype is freely available for ThermoFisher HIV-1 Genotyping Kit users at sanger.exatype.com. We compared the results (FASTA files and patient results) generated by Exatype against our laboratory gold standard method (RECall and Stanford HIVDB. Using a set of 135 sequences, we assessed the proportion successfully analyzed by both methods, as well as the concordance of detection of ambiguous nucleotides, amino acid changes, and drug resistance mutations between the sequences and results generated by the gold standard (Recall and Stanford) and Exatype. While the gold standard produced results for 132 samples, Exatype only produced results for 126. For the 126 (93%) for which results were available from both methods, there was a concordance of 99.8% between the two methods similar to other sequence analysis software comparisons^12,24^. The minor differences were attributed to the partial nucleotide discordance, one method detected a mixture and the other detected one component of the mixture. This consequently resulted in partial discordance for amino acids too.

Exatype and the gold standard had a concordance of 99.1% on NRTI/NNRTI resistance mutations. This is similar to the inter-personnel skill variability on sequence editing^25,26^, depending on the sample tested. The one key resistance mutation mixture that was not detected by the Gold standard and the two that were not detected by Exatype were as a result of partial mismatches due to differential detection of nucleotide mixtures. Despite the high concordance, the inflexibility of a fully automated system may be a drawback to the Exatype system, as the result of this study show in the two key mutations missed. Exatype as the gold standard mark unusual sequence positions, including mixtures, which someone can visually inspect. In our case, Exatype didn’t have any human intervention.

The difference in the numbers of mixtures called between Exatype and the standard method was not statistically significant, making Exatype a vital data analysis standardization tool, especially in clinical reporting, which cannot be achieved by the gold standard method^27–29^. Edits in Exatype are similarly traceable in a separate note pad within the batch system for all the results that are analyzed. This availability makes it compliant to Good Clinical Laboratory Practice (GCLP) standards call for traceability of data in the case of manual edits. Also, Exatype significantly improves the efficiency of HIV drug resistance genotyping and patient reporting. It removes the manual procedure of data transfer and sequence editing that is currently in the gold standard.

The study indicates that the Exatype editing tool had the comparably underestimates the presence of mixtures as opposed to RECall. The discordances in Exatype within the pol sequences were limited to 0.17–1.48% at the NT and AA level, with limited impact on drug resistance interpretations. RECall editing performed slightly better than Exatype editing, as it displayed the highest probability to score mixtures accurately (0.83–1) vs (0.7–0.81). The lowest probability to inaccurately assign mixtures to pure nucleotides (0.002–0.0008). This low probability is attributable to the allowance of sequence editing with RECall. This study also highlighted the necessity of a second inspection as erroneous calls were not only made during automatic but also manual editing. In this respect, Exatype can be made better by allowing sequence editing before result generation.

## Conclusion

RECall editing performed slightly better than Exatype editing, as it displayed the highest probability to score mixtures accurately (0.83–1) vs (0.7– 0.81) and the lowest probability to inaccurately assign mixtures to pure nucleotides (0.002–0.0008). This is attributed to the allowance of sequence editing with Recall. Our results show that Exatype can provide an objective, standardized protocol for HIV sequence analysis for routine patient drug resistance testing and research laboratories, though allowance should be given to allow for sequence editing before result generation for it to be comparable to Recall. The speed and removal of data transfer when using the Exatype is the primary advantage as it removes the sequence edit and the MEGA X QA analysis steps. The system standardizes the laboratory data analysis procedures and thus facilitates unbiased sequence interpretation. We did not factor in the software cost especially for laboratories that are not using the ThermoFischer in-house assay

## Data Availability

Data will be available upon request

## Declarations

### Ethics approval and consent to participate

The study was approved by the Amref Health Africa institutional ethics review committee. This was a retrospective study using remnant viral load samples and the consent of patients to participate was not required.

### Consent for publication

Not applicable

### Availability of data and material

All data contained within the article are publicly available?

### Authors’ contributions

LK conceived the study, collected data, analyzed the data and drafted the manuscript; IM supervised data collection, contributed to data analysis and assisted in drafting and submission of the manuscript. CN, GK, KB, MNK, NB, VO, and DA participated in data collection and review of the manuscript, MK contributed to data analysis, drafting and critical revision of the manuscript. All authors approved the final version of the manuscript.

## Acknowledgments

We acknowledge the Ministry of Health Kenya through the National Public Health Laboratory (NPHL) and the National AIDS and STI Control Program (NASCOP) for facilitating sample collection and allocating staff time to write this manuscript. We thank Peter Young, U.S. Centers for Disease Control and Prevention, Kenya, for reviewing a draft of this manuscript.

